# Spliceosome Inhibition in *SF3B1*-Mutated Uveal Melanoma

**DOI:** 10.1101/2022.09.20.22280164

**Authors:** Josephine Q.N. Nguyen, Wojtek Drabarek, Aïsha M.C.H.J. Leeflang, Tom Brands, Thierry P.P. van den Bosch, Robert M. Verdijk, Harmen J.G. van de Werken, Job van Riet, Dion Paridaens, Annelies de Klein, Erwin Brosens, Emine Kiliç, Rotterdam Ocular Melanoma Study group

## Abstract

Treatment of uveal melanoma (UM) patients with metastatic disease is unfortunately limited. Twenty percent of UM harbor a mutation in splicing factor gene *SF3B1*, suggesting that aberrant spliceosome functioning plays a vital role in tumorigenesis. Splicing inhibitors exploit the preferential sensitivity of spliceosome compromised leukemic cells to these compounds. We have studied the effect of splicing inhibitor E7107 using two UM cell lines and *ex vivo* cultured *SF3B1* and *BAP1* mutated primary UM tumor slices. These UM cell lines and *ex vivo* tumor slices were exposed for 24h to different concentrations of E7107. Tumor slices were stained with H&E and incubated with BAP1, MelanA, MIB-1 and caspase-3 antisera. E7107 exposed UM cell lines showed decreased cell viability and increased apoptosis with the largest effect sizes in *SF3B1*-mutated UM. A similar effect was observed upon exposure of E7107 on UM tumor slices. Additionally, RNA was isolated for transcriptome analysis and the type and number of alternative and aberrant transcripts was evaluated. Ninety-seven transcripts had a decrease in aberrant transcripts in all samples after E7107 exposure, and this effect was mostly in intron retention. This study indicates / suggests that mutated *SF3B1* UM cells are more sensitive to splicing inhibitor E7107 compared to wild-type *SF3B1* UM cells.

**Simple Summary:** Uveal melanoma (UM) is an aggressive malignancy of the eye. UM prognosis varies and depends on mutational status. Mutations in spliceosome gene Splicing Factor 3b subunit 1 (*SF3B1*) are in UM associated with late onset metastasis. Also in other malignancies, such as breast cancer or leukemia, *SF3B1* mutations have been implied as drivers for tumor formation. Targeting the spliceosome with inhibitors is possible and would make *SF3B1*-mutated UM amenable for treatment. We investigated and demonstrated using two cell lines and *ex-vivo* culturing of 3 *SF3B1*-mutated tumors and 17 *SF3B1*-wildtype tumors that spliceosome inhibitor E7107 decreased the cell viability of both *SF3B1*-mutated and *SF3B1*-wildtype UM cell line, but significantly more in *SF3B1*-mutated UM cell line. Therefore, inhibiting the spliceosome has the highest therapeutic potential in *SF3B1*-mutated UM, but further research is recommended to determine the best suited strategy to minimize risk and maximize efficacy in a therapeutic setting.

## 1. Introduction

Uveal melanoma (UM) is the most common primary intraocular malignancy. UM arises from melanocytes in the uvea [1]. In 25-34% of UM patients, tumors metastasize within 10 years, predominantly to the liver, leading to a poor prognosis [2,3]. Prognostic factors include mutations in genes encoding BRCA1 associated protein 1 (*BAP1*) which has the worst prognosis, Splicing factor 3b Subunit 1 (*SF3B1*) with an intermediate prognosis, and Eukaryotic Translation Initiation Factor 1A X-Linked (*EIF1AX*) with the best prognosis[4,5]. Mutations in *SF3B1* occur in approximately 15-25% of UM [6-10] and are associated with early- and late-onset metastasis [11-13]. Furthermore, *SF3B1*-mutated UM have a distinct copy number variation profile compared to other UM tumors, with more aberrations at the distal ends of chromosomes and frequently loss of chromosome 6q and gain of chromosome 6p and 8q [14].

*SF3B1* is, as part of the spliceosome complex, involved in pre-mRNA splicing. Mutations in *SF3B1* result in aberrantly spliced transcripts, which in turn can lead to the formation of aberrant proteins, nonsense-mediated decay (NMD), and downregulation of gene expression [9,15]. An increased sensitivity of a *SF3B1*-mutated leukemia cell line for splicing inhibitors has been observed [16-18]. Furthermore, E7107, one of many spliceosome inhibitors has been studied in phase I trials on human solid tumors [19,20]. E7107 is a first-in-class spliceosome inhibitor, a semi-synthetic derivative of the natural fungal product pladienolide B, which was isolated from Streptomyces platensis [21]. E7107 targets the SF3B subunit 1 and blocks splicing by targeting and binding non-covalently to the SF3B subunit of the U2 snRNP complex. This prevents tight binding of the U2 snRNP complex to the pre-mRNA branch point [22], which results in a decrease of mRNA transcript levels. *SF3B1*-mutated UM cells could be more sensitive to E7107 than *SF3B1*-wildtype cells due to the weakened binding to the branch point [23].

To date, limited treatments are available for metastatic UM patients, including the recently developed protein called Tebentafusp, which has been shown to prolong survival of metastatic UM patients [24]. However, no successful therapy has been developed for specifically *SF3B1*-mutated UM, which be seen as a different subgroup with more similarities to other spliceosome mutated malignancies. By specifically targeting the mutated SF3B1 spliceosome with splicing inhibitors like E7107, tumor progression could potentially be delayed. Therefore, the purpose of this study was to determine both the cytotoxicity and inhibitory effect of E7107 *in vitro* in UM cell lines and *ex vivo* in UM tumors. Due to a less stringent binding of the U2 snRNP complex to the branch point, we hypothesize that the susceptibility of *SF3B1-*mutated UM to E7107 is greater than that of *SF3B1*-wildtype UM. Hence, E7107 or similar compounds could have potential as a treatment modality for *SF3B1*-mutated metastatic UM.

## 2. Materials and Methods

### 2.1. Cell culture

Two established UM cell lines (Mel202 and 92.1) were used for this study (Mel202 kindly gifted by dr. B Ksander, Schepens Eye Research Institute, Harvard, Boston, MA, USA and 92.1 kindly gifted by dr. I de Waard-Sieblinga, Leiden University, Leiden, The Netherlands). Mel202 and 92.1 cells were cultured in RPMI-1640 medium containing L-glutamine (Gibco, Thermo Fisher Scientific, Waltham, MA, USA), supplemented with 10% Fetal Calf Serum (FCS; Biowest, Nuaille, France) and 2% penicillin/streptomycin (P/S; Gibco, Thermo Fisher Scientific, Waltham, MA, USA). Mel202 harbors a *SF3B1* mutation (*SF3B1*^MUT^) c.1793C>T (R625G) and 92.1 harbors an *EIF1AX* mutation (*SF3B1*^WT^) c.17G>A (G6D) (both have been verified using Sanger Sequencing) [25]. Cells were grown at 37°C in a 5% CO2 humidified atmosphere (Heracell 150 CO2 incubator, Heraeus, Hanau, Germany).

### 2.2. Cell viability and density analysis

To determine the cytotoxic effect of E7107, the cell viability of *SF3B1*^MUT^ and *SF3B1*^WT^ UM cells was assessed after treatment with different concentrations of E7107 (gifted by H3 Biomedicine Inc, Cambridge, MA, USA). To evaluate the toxicity effects of DMSO as solvent, we also used a negative control (0 nM E7107) to assess the solvent toxicity. *SF3B1*^MUT^ and *SF3B1*^WT^ UM cell line were seeded (2.0 × 10^5^ per well in 6 wells-plate) for 24 hours before exposure for 24 hours to E7107 (concentration range 0-5, 7, 10, and 20 nM E7107). The cell viability was assessed by counting the number of viable cells per condition in triplicate with trypan blue dye (#1450021, Bio-Rad, Hercules, Ca, USA) using the TC20TM Automated Cell Counter (Bio-Rad, Hercules, CA, USA). To determine the timing of the cytostatic effect of E7107, the cell density of *SF3B1*^MUT^ and *SF3B1*^WT^ UM cell line was assessed during exposure to E7107 of 24 hours of incubation. This was assessed in two different time-lapse experiments. In the first experiment, cells (5.0 × 10^3^ per well in black 96 wells-plate) treated with E7107 (0-20 nM) were imaged on the Opera Phenix High Content Screening System (PerkinElmer, Waltham, MA, USA) using the 10x air objective (0.3 NA) for a period of 21 hours with a time interval of 3 hours. In the second experiment, cells (2.0 × 10^3^ per well in black 96 wells-plate) were treated with E7107 (0-0.5 nM) and images were taken for a period of 69 hours with a time interval of 3 hours for the first 24 hours, and afterward with a time interval of 6 hours. The captured images were analyzed with the associated Harmony office software (PerkinElmer, Waltham, MA, USA). The software calculated the percentage decrease or increase in cell area compared to 0h after each interval.

### 2.3. Tumor tissue work up

Fresh UM tissue was collected from UM patients undergoing enucleation at the Erasmus MC University MC Rotterdam and the Rotterdam Eye Hospital Rotterdam, The Netherlands from September 2018 - September 2021. Twenty tumor tissues were collected at the Department of Pathology. Iris melanoma and irradiated tumors were excluded from this study. Mutational status was determined by BAP1 immunohistochemistry, Sanger sequencing and/or Next Generation sequencing [26,27]. The tissues were embedded in 4% agarose low gelling temperature (#A9414-100G, Sigma Aldrich, Saint Louis, Mo, USA) at 37°C before automated tissue slicing using a Leica VT 1200S Vibratome (Leica Biosystems Inc., Buffalo Grove, IL, USA) with 0.8 mm/s slicing speed, 300 μm slice thickness, and 2.0 mm vibration amplitude. Slices were cultured in 6-wells plate in culture medium (RPMI-1640 medium containing L-glutamine (Gibco, Thermo Fisher Scientific, Waltham, MA, USA), supplemented with 10% FCS (Biowest, Nuaille, France) and 2% penicillin/streptomycin (P/S; Gibco, Thermo Fisher Scientific, Waltham, MA, USA) and supplemented with either 0, 1, and 5 nM E7107 or solvent control at 37°C for 24 hours on a Stuart SSM1 mini orbital shaker (Cole-Parmer, Staffordshire, UK) at 60 rpm. Afterward, slices were used for either RNA isolation or fixation for immunohistochemistry (IHC). The quantity and quality of the RNA was determined using the Bioanalyzer (Agilent Genomics, Santa Clara, CA, USA). Tumors where a full set (medium, 0, 1 and 5 nM E7107) of RNA is available with a high RNA quality (RIN > 8.0) were eligible for RNA sequencing.

### 2.4. Reverse transcription polymerase chain reaction (RT-PCR)

RNA isolation was performed using the miRNeasy Mini kit (Qiagen, Hilden, Germany) according to the manufacturer’s protocol and cDNA was synthesized with 1 μg RNA using the iScript cDNA Synthesis Kit (Bio-Rad Laboratories, Veenendaal, The Netherlands) according to the manufacturer’s protocol. Custom primers on the exons adjacent to the splicing aberration were designed using primer3 (Table S1) for RT-PCR (Biometra TAdvanced PCR Thermocycler) for five candidate genes Armadillo Repeat Containing 9 (*ARMC9*), Enolase Superfamily Member 1 (*ENOSF1*), Diphthamide Biosynthesis 5 (*DPH5*), Dihydrolipoamide S-Succinyltransferase (*DLST*), and Cyclin-dependent kinase 2 (*CDK2*). Four of these (*ARMC9, ENOSF1, DPH5* and *DLST*) have proven to be aberrantly spliced in *SF3B1*^MUT^ UM cell lines [9]. After re-analyzing the UM RNA-seq data of Wojtek et al.[13], we included one additional gene *CDK2*, and two control genes Charged Multivesicular Body Protein 2A (*CHMP2A*) and ER Membrane Protein Complex Subunit 7 (*EMC7*). Results were analyzed using the image analysis software ImageQuantTL to determine the effect of E7107 on splicing inhibition in aberrant transcripts and canonical transcripts. Here, transcripts were defined as “canonical” when speaking of unaffected transcript lengths of known isoforms. Linear regression analysis was used to determine the degree of splicing inhibition evoked by E7107 by calculating the slope.

### 2.5. RNA sequencing and analysis

First, strand-specific cDNA libraries were generated with the strand-specific NEBNext Ultra II Directional RNA Library Prep Kit protocol and polyA mRNA workflow (NEB #E7760S/L) on Illumina NovaSeq6000 (Illumina, San Diego, USA). The sample preparation was performed according to manufacturer’s protocol (GenomeScan, Leiden, The Netherlands). Quality control and read trimming were performed with Trim Galore v.0.6.7 [28]. Read alignment, transcript quantification and differential expression analysis were performed using CLC Genomics Workbench version 20 (Qiagen, Hilden, Germany). Reads were aligned to the human reference genome (hg_g1k_v37) according to the following settings: mismatch cost 2, insertion/deletion cost 3, length fraction 0.8, similarity fraction 0.8, alignment to gene regions only. Transcripts including mitochondrial, ribosomal RNA and microRNA were excluded in the analysis. Paired reads were counted as one. Alignment metrics can be found in the supplementary (Table S2, Figure S1). Trimmed mean per million (TMM) values was used to normalize for sequencing depth across samples. For each gene, counts per million (CPM) and transcripts per million (TPM) were calculated. Total read counts can be found in the supplementary (Table S3).

MISO (Mixture-of-Isoforms) v.0.5.4 was used to determine and compare the number of alternatively spliced genes due to differential expressions of isoforms and exons across samples [29]. Reads are aligned to the hg19 reference using HISAT2 (hg_g1k_v37) and exon-isoforms distributions for each sample are compared by counting and normalizing known isoforms-exon usage. For event selection, the results were filtered for a deltaPsi (ΔΨ) ≥ 0.1 and a Bayes factor of at least 10. In addition, FRASER (Find RAre Splicing Events in RNA_seq) determined potential candidates for aberrant splicing events on a cohort level [30]. Here, the same ΔΨ cut-off was applied as expression filtering. For the computation of the p- and z-values, ψ5 (alternative acceptors) = 3, ψ3 (alternative donors) = 5 and θ (splicing efficiencies) = 2 were used. The results for individual samples were ranked according to their ΔΨ values and the top 25 genes of each sample were combined for comparison based on their psi/theta scores. Afterwards, candidates were included when a decrease in the ΔΨ value was assessed per set, since this showed that a decrease in aberrant transcripts formation occurs when treated with E7107.

The change in Copy Number Variation (CNV) profiles and allelic distributions of the tumors treated with E7107 were estimated using SuperFreq [31,32]. BAM files were available from the HISAT2 mapping and VCF files were made according to VarScan (variant detection *p*-value of 0.01, variant frequency of at least 0.02 and no strand filter). SAMtools mpileup was used with a maximum depth of 1000 reads to compensate for highly covered regions, a minimum base and mapping quality of 15 and the same reference genome as for the HISAT2 alignment. The results of SuperFreq were obtained using the default settings of the RNA mode along with the hg19 genome (hg_g1k_v37). Samples were normalized against the ‘medium only’ condition.

### 2.6. Immunohistochemistry (IHC)

After formalin fixation, tumor slices were embedded in paraffin and 4 μm sections were generated for microscopy analysis using the microtome. Immunohistochemistry (IHC) was performed with an automated, validated, and accredited staining system (Ventana Benchmark ULTRA, Ventana Medical Systems, Tucson, AZ, USA) using ultraview or optiview Universal Alkaline Phosphatase Red Detection Kit (#760-501). Following deparaffinization and heat-induced antigen retrieval, the tissue samples were incubated according to their optimized time with the antibody of interest (Table S4). Incubation was followed by hematoxylin II counter stain for 12 minutes and then a blue coloring reagent for 8 minutes to turn the purple hematoxylin blue according to the manufactures instructions (Ventana Benchmark ULTRA, Ventana Medical Systems, Tucson, AZ, USA). We used hematoxylin and eosin (H&E) to examine the histological tumor architecture IHC for melanoma antigen (MelanA) was performed to identify areas with >90% tumor cells, and BAP1 [26] immunostaining to determine whether tumors are BAP1 positive or negative, thereby indicating if a *BAP1* mutation is present. Immunohistochemical detection of the proliferation marker Ki67 was performed using the monoclonal antibody mind bomb E3 ubiquitin protein ligase 1 (MIB-1), evaluating areas containing >90% tumor cells. Similarly, caspase-3 immunostaining was used to detect apoptotic cells.

### 2.7. Statistical analysis

Statistical analysis was performed using GraphPad Prism 9.0 (GraphPad Software, Inc., San Diego, CA, USA). Two-way ANOVA with Tukey’s test or paired Student’s t-test were used to compare differences between groups and concentrations. Linear regression was performed to determine the degree of splicing inhibition. The LC (Lethal Concentration) 50 value was defined as the concentration which cell viability decreased with 50%. The mean positive cells detected with IHC on slices was calculated per mm^2^. When slices were smaller than 1 mm^2^, the mean of the positive cells was scaled to the correct size for the analysis. A *p-*value of < 0.05 was considered statistically significant.

## 3. Results

### 3.1. Cytotoxicity and proliferation inhibition of E7107 in vitro

The cytotoxicity of E7107 was assessed in a single-dose administration in Mel202 *SF3B1*^MUT^ and 92.1 *SF3B1*^WT^ cells. A decrease in cell viability was observed when E7107 was administered in a dose-dependent manner for both cell lines (Figure 1). The decline in cell viability was statistically significant after treatment with 2 nM E7107 and onwards compared to medium and 0 nM E7107 in both cell lines (*p* < 0.05). Furthermore, the toxicity was significantly higher in Mel202 *SF3B1*^MUT^ cells (*p* = 0.0004). The LC50 values were 2.66 nM and 5.67 nM in Mel202 *SF3B1*^MUT^ and 92.1 *SF3B1*^WT^ UM cell lines respectively, with a cytotoxic potency of E7107 2.1 times higher in Mel202 *SF3B1*^MUT^ cells than in 92.1 *SF3B1*^WT^ cells. The proliferation was inhibited in both Mel202 *SF3B1*^MUT^ and 92.1 *SF3B1*^WT^ UM cells after treatment with E7107 in a time-and dose-dependent manner (Figure 2). In both cell lines, DMSO (0 nM E7107) seemed to have a positive effect on proliferation. Cells were still proliferative until 0.5 – 1 nM E7107, and the proliferation stopped. A significant decrease in cell proliferation was assessed after 21 hours in *SF3B1*^MUT^ cells with a concentration of 10 nM E7107 and above (*p* = 0.0007) whereas only a concentration of 15 nM E7107 was significant in *SF3B1*^WT^ cells (*p* = 0.0089). After 69 hours, no significantly differences were observed in Mel202 *SF3B1*^MUT^ cells while 92.1 SF3B1^WT^ cells were significantly inhibited with a concentration of 0.2 and 0.5 nM E7107 (*p* = 0.0019).

**Figure 1.**
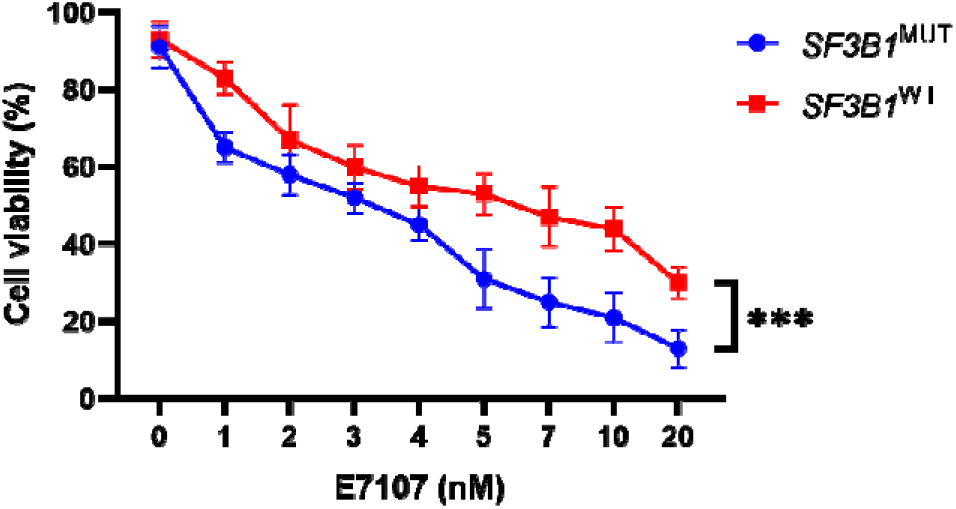
The effects of E7107 to cell viability in uveal melanoma cell lines. Mel202 *SF3B1*^MUT^ and 92.1 *SF3B1*^WT^ UM cells were treated with 0-20 nM E7107 for 24 hours. Paired Student’s t-test and two-way ANOVA was used to calculate statistical significance between both cell lines and between concentrations, respectively. A statistical significance between both cell lines was observed in all concentrations. When compared to medium and to 0 nM E7107, a concentration of 2 nM E7107 and onwards was statistically significant in both cell lines. *** = *p* < 0.001; (N>3).

**Figure 2.**
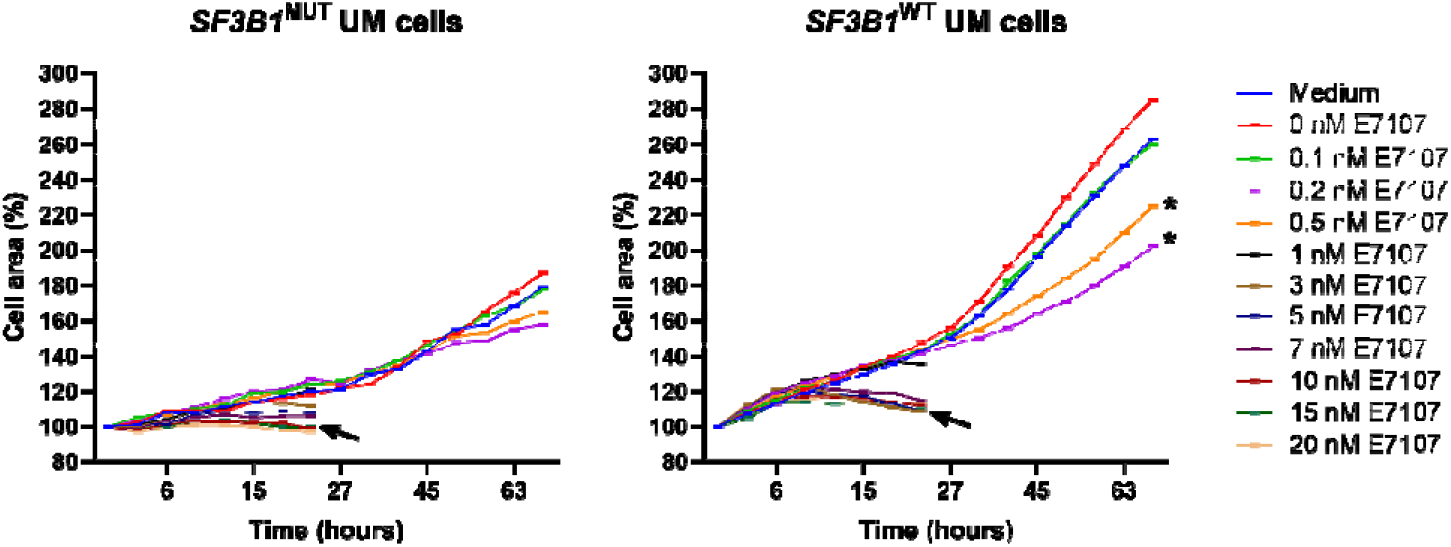
Growth rate inhibition after exposure to E7107. Drug potency was determined on *SF3B1*^MUT^ and *SF3B1*^WT^ UM cells using Opera Phenix High Content Screening System. A significant decrease in proliferation inhibition was observed after 21 hours with a concentration of 10 nM E7107 (arrow) and above (*p* = 0.0007) in *SF3B1*^MUT^ cells and 15 nM E7107 (arrow) in *SF3B1*^WT^ cells. After 69 hours, no significantly differences were observed in *SF3B1*^MUT^ cells while a concentration of 0.2 and 0.5 nM E7107 (asterisk) was significant in *SF3B1*^WT^ cells (*p* = 0.0019).

### 3.2. Transcription level analysis

The sensitivity of E7107 on splicing inhibition was assessed on five target genes of interest (*ARMC9, ENOSF1*, DPH5, DLST and *CDK2*) and two control genes (*CHMP2A* and *EMC7*) in both SF3B1^MUT^ and SF3B1^WT^ cell lines. Due to the mutation in SF3B1, aberrant transcripts are present in Mel202 SF3B1^MUT^ cells. Both canonical and aberrant spliced mRNA transcripts with extended exons were assessed in Mel202 SF3B1^MUT^ cells, whereas only canonical transcripts were observed in 92.1 SF3B1^WT^ cells (Figure 3a, S2). In general, treatment with E7107 inhibited splicing in a dose-dependent manner in both cell lines based on the product intensity. Both the canonical and the aberrant transcipts were affected, yet a stronger decline of the amount of aberrant, compared to canonical, transcripts from the genes-of-interests (using linear regression) was observed within Mel202 SF3B1^MUT^ cells (Figure S3). For example, the linear regression of aberrant transcripts was −0.1091 compared −0.07337 (*p* = 0.1195) in the canonical transcripts for *ARMC9*.

**Figure 3.**
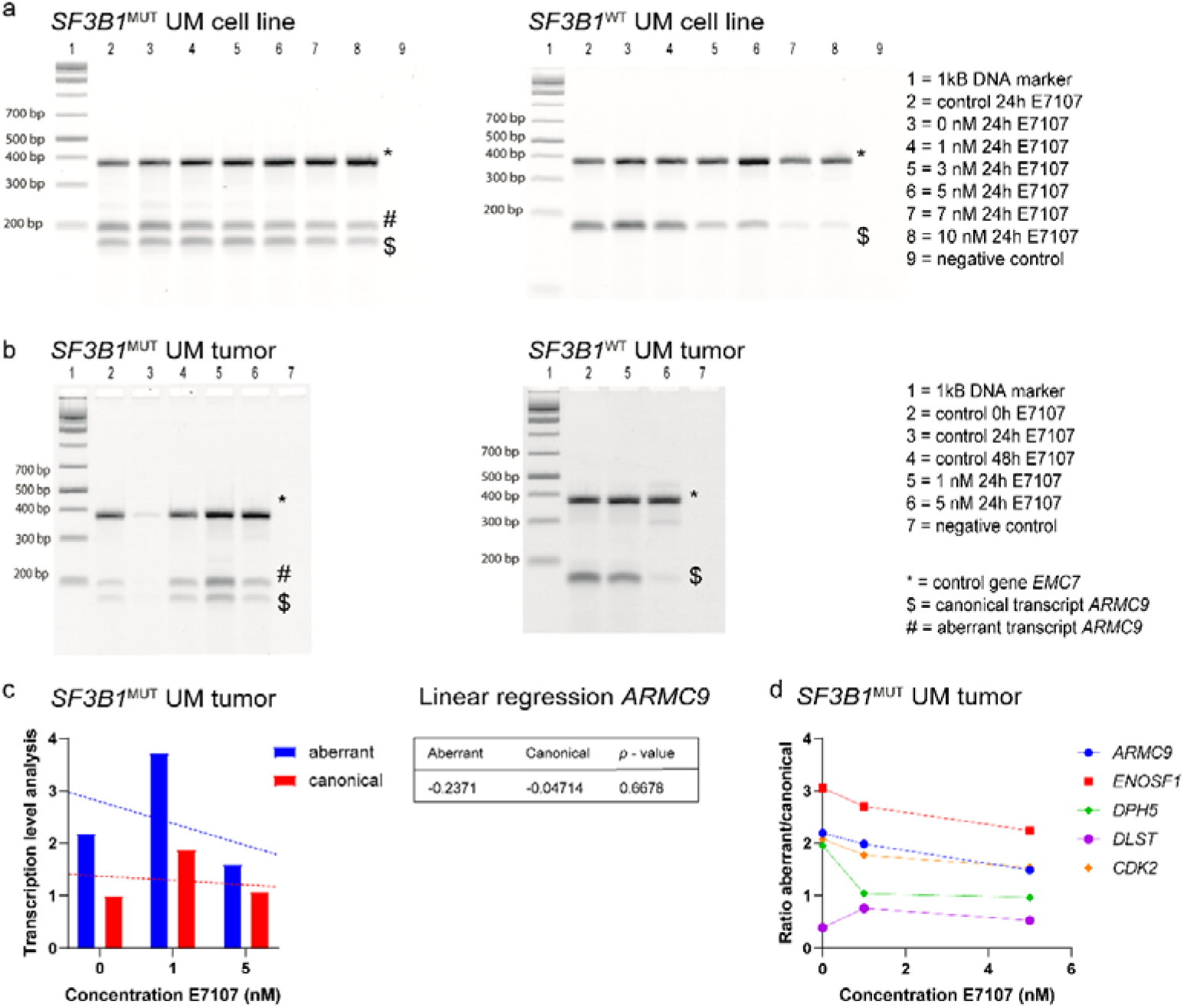
RT-PCR analysis of the effect of E7107 treatment on splicing inhibition on UM cell lines and tumors. Multiplex RT-PCR products for *ARMC9* on Mel202 *SF3B1*^MUT^ and 92.1 *SF3B1*^WT^ cells (a) and tumors (b) treated with different concentrations of E7107. Amplification of the *SF3B1*^MUT^ sensitive genes resulted in both aberrant and canonical transcript formation for both *SF3B1*^MUT^ cells and tumor, whereas the *SF3B1*^WT^ products only displayed normal transcript formation. (c) Aberrant and canonical transcripts formation was quantified using ImageQuantTL in *SF3B1*^MUT^ tumor on *ARMC9*. Linear regression analysis was used to determine the degree of splicing inhibition. (d) The ratio of aberrant versus canonical transcripts for various genes in a *SF3B1*^MUT^ tumor (tumor 01). The insensitive gene *EMC7* was used as a control for both cDNA quality and quantity. *SF3B1*^WT^ tumor had an *EIF1AX* mutation. The analysis of other genes can be found in the supplementary (Figure S3).

### 3.3. Tumor tissue

Fresh UM tissue of twenty patients undergoing enucleation was collected and mutational status of key UM-related hotspot genes for all tumors was determined (Table S5). Seventeen tumors harbored a primary driver mutation in G protein Subunit Alpha Q (*GNAQ*) or G protein Subunit Alpha 11 (*GNA11*), in one tumor a mutation in Cysteinyl Leukotriene Receptor 2 (*CYSLTR2*) and in one other a mutation in Phospholipase C Beta 4 (*PLCB4*) was observed. In almost all tumors a secondary driver mutation was detected: ten patients harbored a mutation in *BAP1* and or were BAP IHC negative, three in *SF3B1* and five in *EIF1AX*.

### 3.4. Splicing inhibition assay ex vivo

The splicing inhibitory effect of E7107 on UM tissue slices was assessed with RT-PCR using the same genes as within our *in vitro* experiments. The formation of both canonical and aberrant transcripts was observed in *SF3B1*^MUT^ tumor (tumor 01) whereas *SF3B1*^WT^ tumor (tumor 03) had only canonical transcript formation (Figure 3b). The RT-PCR analysis revealed that mRNA levels of these candidate genes decreased, both in the canonical transcripts as well as the aberrant transcripts. These observations were dose dependent (Figure 3c). Furthermore, we have found a decrease in the ratio aberrant versus canonical in these genes of interest, especially at a concentration of 5 nM E7107 (Figure 3d).

### 3.5. RNA sequencing and splicing analysis

To validate our findings of the selected genes, we have evaluated the inhibitory effects transcriptome wide. RNA sequencing was performed on two cell lines (*SF3B*1^MUT^ and *SF3B1*^WT^) and two tumors (*SF3B1*^MUT^ [tumor 14] and *SF3B1*^WT^ [tumor 06]). We evaluated the transcriptomes of these four samples exposed to: medium, 0, 1 and 5 nM E7107. We have assessed the use of alternative transcripts usage with the MISO analysis after treatment with E7107. An increase in known isoforms usage was seen in all samples, with the largest effect in *SF3B1*^MUT^ cells with a concentration of 5 nM E7107. However, no significant differences were observed in alternative transcript (known isoforms) usage between the different concentrations E7107 (Figure 4, Table S6). Next, the FRASER analysis was used to assess aberrant splicing events. Aberrant splicing was categorized in three event groups: intron retention (θ), aberrant donor site usage (ψ3) and aberrant acceptor site usage (ψ5). Overall, no striking differences were observed between samples and between conditions (Figure S4). It seems like cell lines have lower events in general compared to tumor slices, however more samples are required for a statistical analysis. When looking at the top 25 genes of each sample and combining these, ninety-seven transcripts had a decrease in aberrant transcripts formation after exposure to E7107 (Table S7). Most of these events were intron retention, followed by aberrant acceptor site usage and aberrant donor site usage (Figure 5). Furthermore, the Superfreq CNV and allelic distribution estimates hint at an increasing effect of E7107 (Figure 6, S4). However, these estimates could also be due to specific high cell clonality, which could be more present in a large fraction of the test samples. To assess the proliferation and viability effects on protein level, we next detected positive cells on tumor slices with IHC.

**Figure 4.**
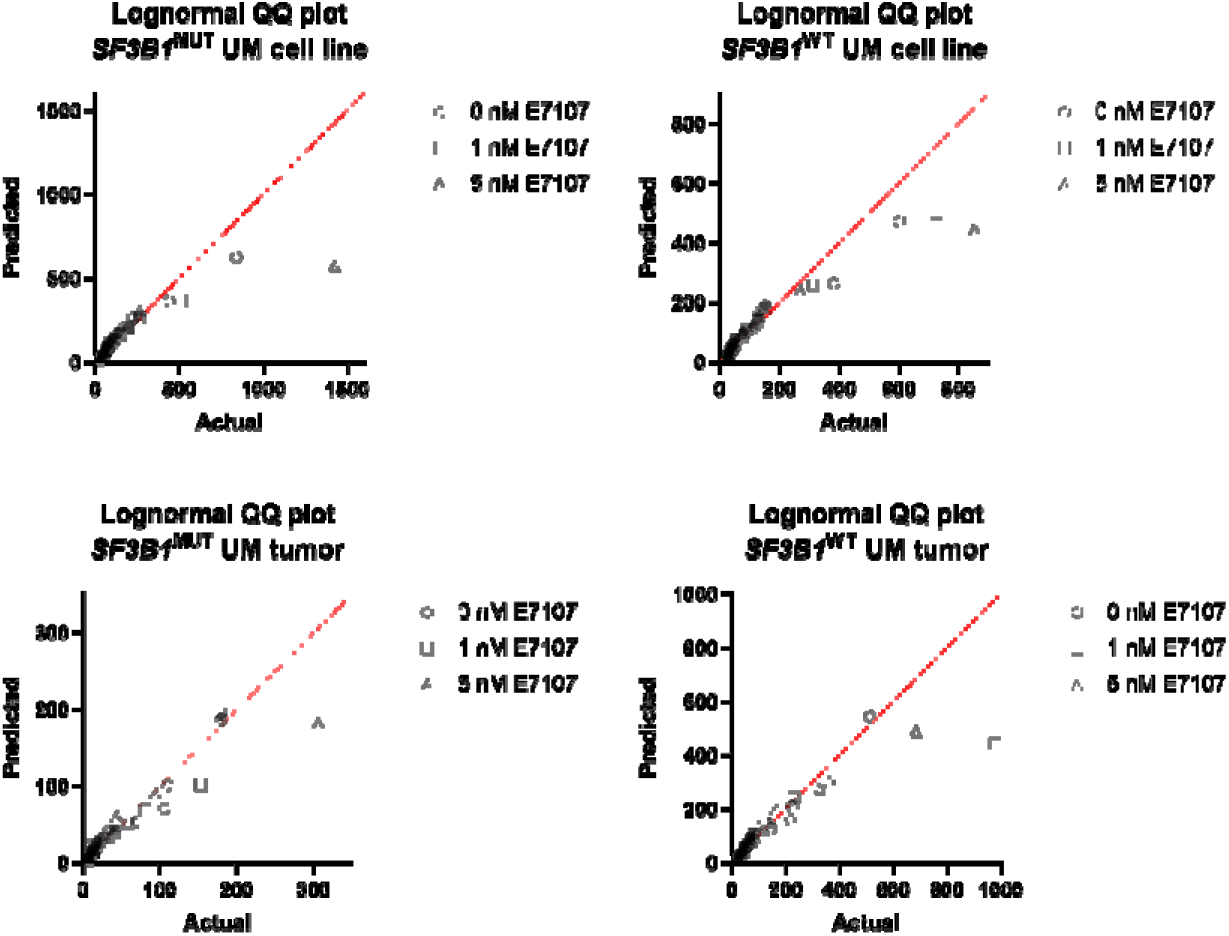
Normal distribution Q-Q plots of alternative splicing. Q-Q plots of the samples with the E7107 concentrations. No differences were observed between a normal distribution and the distribution of alternative splicing usage with the Kolmogorov-Smirnov test (*p* > 0.05, Table S7).

**Figure 5.**
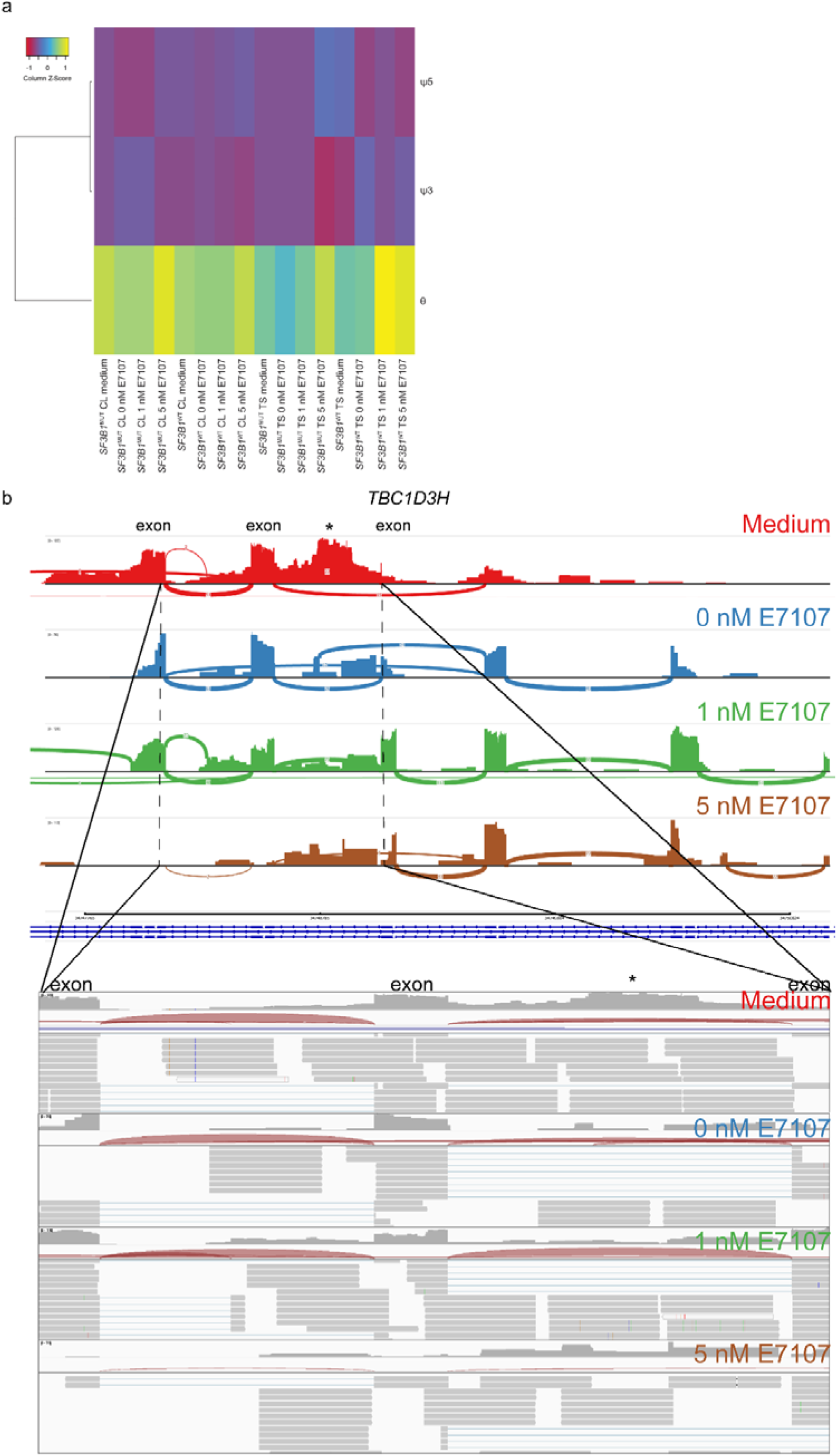
Aberrant splicing analysis. (a) Heatmap of aberrantly spliced genes per sample per event: intron retention (θ), aberrant donor site usage (ψ3) and aberrant acceptor site usage (ψ5). A higher number of intron retention was observed compared to aberrant donor/acceptor site usage events. The intron retentions were most often seen in 5 nM E7107 samples. CL, cell line; TS, tissue slice. (b) Sashimi plot of the intron retention event (*) in the *SF3B1*^MUT^ tumor RNA-seq samples of gene TBC1 Domain Family Member 3H (*TBC1D3H*) as one of the top genes as example. A decrease in intron retention is observed, mostly in 5 nM E7107.

**Figure 6.**
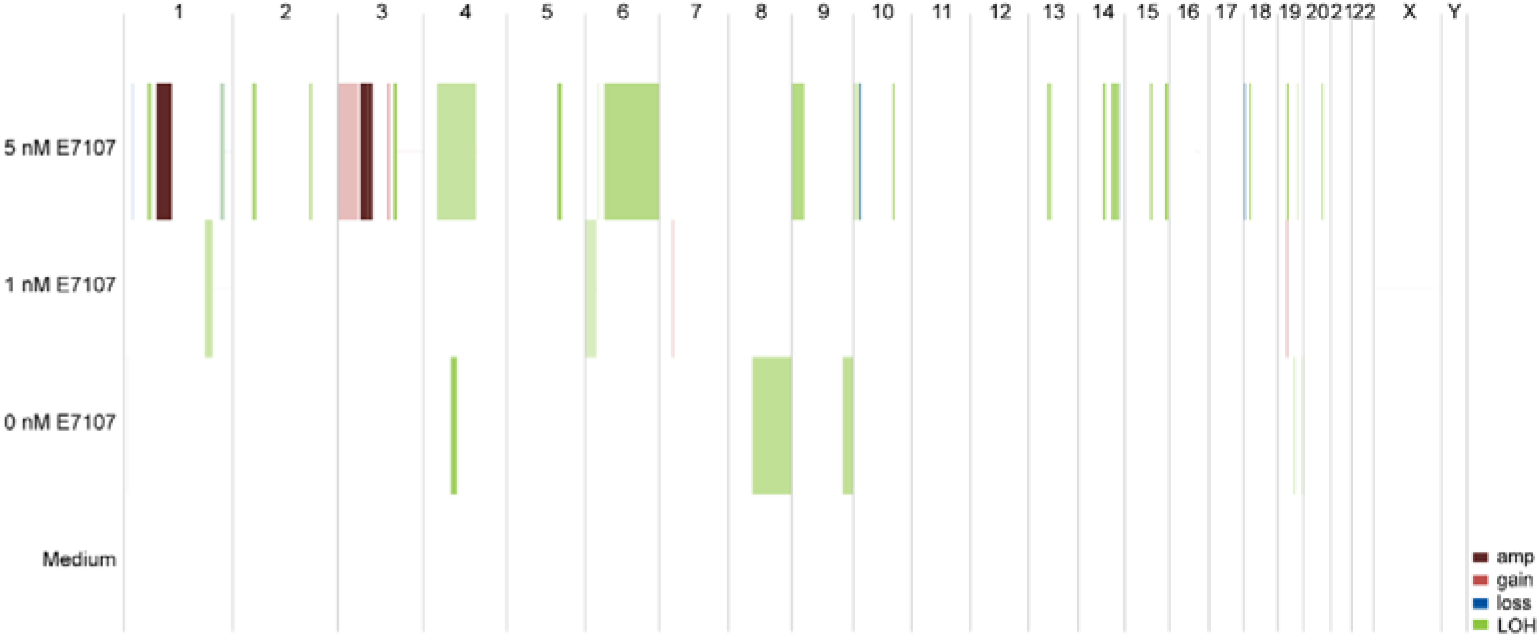
Estimates of the effect of increasing E7107 concentrations on zygosity and relative DNA copy number of the *SF3B1*^MUT^ tumor slices (tumor 14) per condition from whole transcriptome data. The Superfreq DNA copy number and allelic distribution estimates hint at an increasing effect of E7107. An increase in copy number neutral loss of heterozygosity (LOH) is seen after E7107 exposure, especially at 5 nM E7107, as well as an increase in allele copies. Amp, 4+ copies; gain, 3 copies; loss, <2 copies; LOH, 2 copies of the same allele. Each chromosome is depicted on the x-axis. The conditions were normalized to medium. The tumor CNV can be found in the supplementary (Figure S5b-c).

### 3.6. IHC staining

The morphology and detection of tumor cells were assessed with H&E and Melan-A staining, respectively. More apoptotic cells were seen with E7107 treatment. There was a general trend towards an increase in apoptotic cells (caspase-3) in all tumor slices, with the biggest increase in *SF3B1*^MUT^ tumor slices treated with 5 nM E7107 (Figure 7). A slight increase in proliferation (MIB-1) was assessed in all samples (*p* = 0.4958).

**Figure 7.**
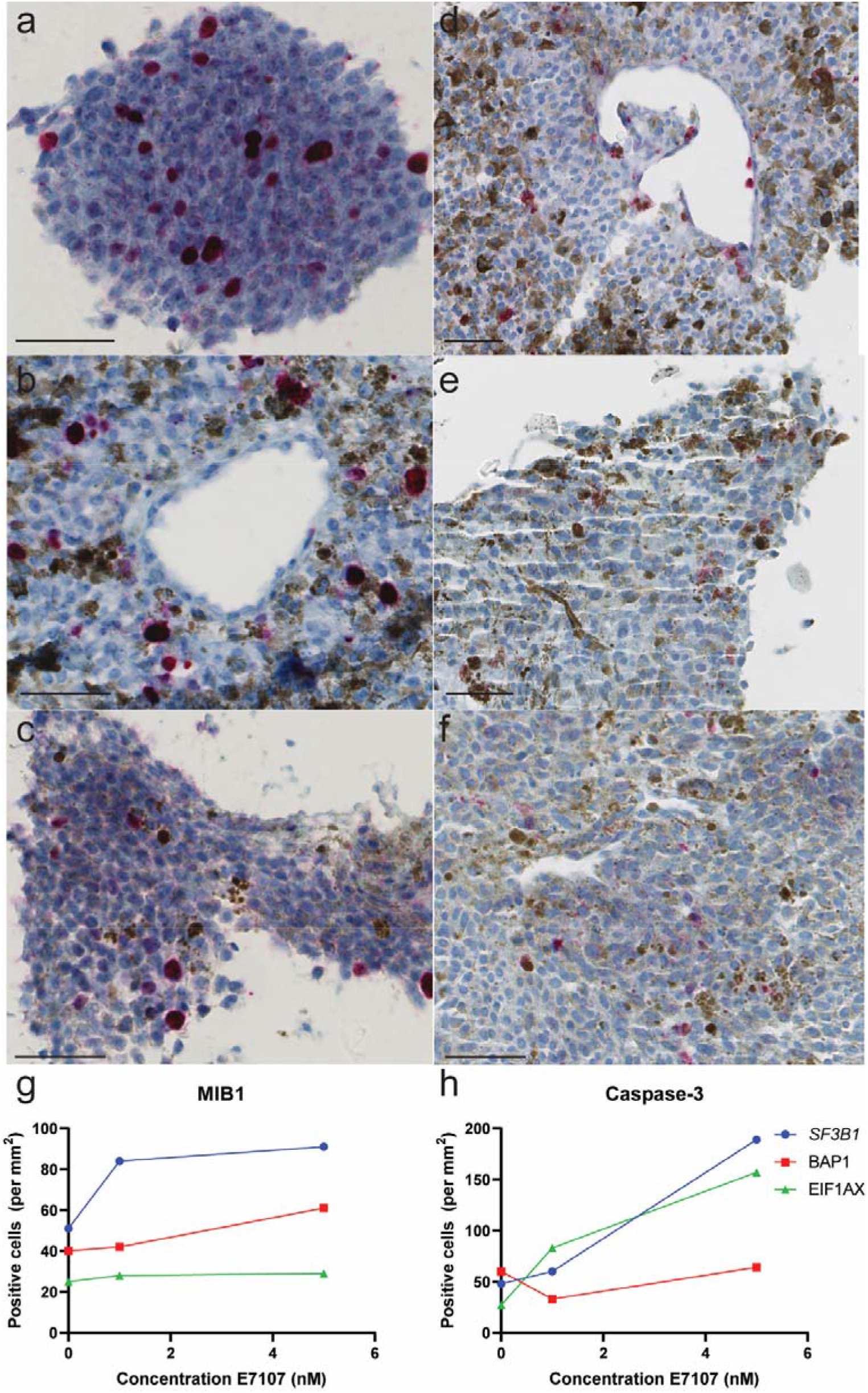
Immunohistochemistry staining of UM tumor tissues treated with 5 nM E7107. Proliferating cells were marked with mind bomb E3 Ubiquitin Protein Ligase 1 (MIB-1) in (a) *SF3B1*^MUT^ tumor (tumor 01), (b) *BAP1*^MUT^ tumor (tumor 19) and (c) *EIF1AX*^MUT^ tumor (tumor 03). An increase in apoptotic cells was seen in (d) *SF3B1*^MUT^ tumor (tumor 14), (e) *BAP1*^MUT^ tumor (tumor 02) and (f) *EIF1AX*^MUT^ tumor (tumor 03). with caspase-3 antibody (scale bar 50 μm). The mean positive cells per mm^2^ is visualized per mutation for MIB-1 and caspase-3 (g,h).

## 4. Discussion

In this study, we assessed the cytotoxicity and splicing inhibitory effects of E7107 *in vitro* on UM cell lines and *ex vivo* on UM tumor tissue slices. While splicing is an essential process in all mammalian cells, this study suggests that the viability of *SF3B1*^MUT^ UM is reduced to splicing inhibitor E7107 compared to *SF3B1*^WT^ UM.

Our results suggest that E7107 decreases splicing of mRNA transcripts more in *SF3B1*^MUT^ cells compared to *SF3B1*^WT^ cells in a dose-dependent manner with an optimal concentration of 5 nM E7107 with RT-PCR. In our *in vitro* proliferation analysis, no significant differences were observed after 69 hours in Mel202 *SF3B1*^MUT^ cells, whereas 92.1 *SF3B1*^WT^ cells showed significant inhibition with 0.2 and 0.5 nM E7107. An explanation for these skewed results could be that E7107 is more toxic in Mel202 *SF3B1*^MUT^ cells where apoptosis occur earlier in the affected cells, and therefore the results were mainly of the unaffected cells. On the contrary, 92.1 *SF3B1*^WT^ cells could be less affected by E7107 where these cells need more exposure time to reach the same effect. Furthermore, the two cell lines used in this study (Mel202 and 92.1) have a different proliferation rate and doubling time, hence making it difficult to interpret and compare results [25]. Moreover, the transcriptome-wide aberrant splicing analysis showed overall high aberrant donor/acceptor site events in all samples. Nonetheless, when looking at the effects of E7107 exposure on the aberrant splicing events per transcript, a decrease in aberrant splicing events were mainly in intron retention events. Therefore, E7107 has the largest effect on intron retention, consistent with literature [17]. Since there were no biological replicates in the RNA-seq analysis due to limited material, including more samples and/or including single-cell RNA seq data in the future would strengthen the analysis. With the CNV analysis on RNA-seq data, it seems to indicate that there is loss of a specific allele with compensation of one extra identical copy after E7107 exposure. An explanation for this could be that only subclones were analyzed due to the toxicity of E7107 (cell viability assay). However, it remains difficult to conclude this based on RNA-seq data. Furthermore, we have confirmed our findings on the viability on protein level with IHC. An increase in apoptotic cells was seen with E7107 treatment in all samples. Apoptosis in lower concentrations could also be a result of the toxicity of the solvent DMSO, as DMSO can be toxic to cells in certain conditions (such as exposure for prolonged periods or when high concentrations are used) [33]. A slight increase in proliferating cells was observed in all tumors after E7107 exposure. In general, approximately 2% of UM cells are MIB-1 positive, hence this increase in positive cells could also be due to non-specific staining of lymphocytes [34]. Even though the analyzed areas were selected with > 90% tumor cells, infiltrating lymphocytes in the tumor could not always be excluded.

The inherent sensitivity of mutated cells to E7107 and other splicing inhibitors has also been assessed in other studies [35-39]. Obeng et al assessed an increased sensitivity to E7107 in *SF3B1*^MUT^ (K700E) murine myelodysplastic syndrome (MDS) cells *in vitro* and *in vivo* [18]. Similar to our results, Obeng et al. showed that *SF3B1*^MUT^ murine MDS cells are more sensitive to E7107 (cell viability = 0.619 nM) than *SF3B1*^WT^ cells (cell viability = 1.249 nM), strengthening our findings. Notably, these low values show a strikingly smaller toxicity range compared to our values of 2.66 nM in Mel202 *SF3B1*^MUT^ and 5.67 nM in 92.1 *SF3B1*^WT^ cell lines. Mice were injected E7107 daily intravenously in the tail vein with a dose of 4 mg/kg for 10 days (5 days on, 2 days off, then repeat*). In vivo* results indicated a similar increased sensitivity to E7107 of the mutated cells compared to wild-type cells. Furthermore, the effects of E7107 have also been evaluated in *SRSF2*, another splicing gene [17]. Treatment of E7107 in murine myeloid leukemia cells resulted in preferential cell death of *SRSF2*-mutated cells, with a dose of 4 mg/kg intravenously for 10 consecutive days, similar to the study mentioned above. Even though mutation in *SRSF2* have been detected in UM, it occurs less frequently compared to leukemia [40,41]. Nonetheless, E7107 only exerts its splicing inhibitory effects by binding directly to the SF3b complex. A previous identified mutation in *SF3B1* (R1074H) has been shown to be less sensitive to E7107 due to its inability to bind to the SF3b complex [42]. Lee et al. confirmed these findings with a 300-fold greater IC^50^ in *SF3B1*^R1074H^ cells compared to *SF3B1*^WT^ cells [17]. Even though mutation in *SF3B1* at location R1074H have not been reported before in UM [43], this underlines the importance of the specificity of E7107 in spliceosome mutant malignancies.

Here, our analysis showed a clear inhibition of cell proliferation until 21 hours. After that period of time, the effects of E7107 seemed less effective regardless of the mutation status. Furthermore, the LC50 values in our study are higher compared to other studies [17,18]. The values were higher for both *SF3B1*^MUT^ and *SF3B1*^WT^ cells. This could be due to the difference in cell type (human vs murine) or E7107 treatment (24 hours vs 72 hours). Another explanation could be that uveal melanoma cells are less sensitive to E7107 *in vitro* compared to other malignant cells such as MDS. The *in vitro* concentration range of E7107 in our study is within range of other studies [17,18,44]. Furthermore, a plateau phase was observed between 0.5 and 1 nM E7107 in our proliferation assay, which coincide with the LC50 values [0.619 – 0.90 nM] of these studies [17,18]. Even though the LC50 values of these studies varied in cell lines, they used the same dose for *in vivo* (4 mg/kg/day) experiments. With this dose, no evident toxicity of E7107 was mentioned in these studies. However, in contrast to UM which has a doubling time of 154-511 days (63 days in metastatic UM), murine leukemic cells have a doubling time of only approximately 0.5 days [45-47].

To date, E7107 has been studied in two separate phase I clinical trials in patients with solid tumors. In both trials, E7107 reversibly inhibited pre-mRNA in a dose-dependent manner at the maximum tolerated dose of 4.0 – 4.3 mg/m^2^ [19,20]. E7107 was administered intravenously, with a bolus of 5 to 30 minutes on day 1, 8 and 15 [19] and on day 1 and 8 [20]. In both studies, E7107 had a high systemic clearance, with a comparable plasma elimination half-life (Eskens et al.: 5.3 – 15.1 hours and Hong et al.: 6 – 13 hours). Furthermore, after discontinuation of E7107 administration, a stable disease was observed with no changes in the size of metastases after follow-up. E7107 was generally well tolerated, with the most common drug-related adverse events being gastrointestinal related such as nausea and vomiting. However, visual impairment was reported in 3 out of 66 patients and this resulted in termination of both trials. Since our study evaluated the inhibition effects *in vitro* and *ex vivo*, this toxicity could not be assessed. It remains unclear if the observed toxicity in both trials is due to *SF3B1* inhibitors in general or specifically to E7107. Nonetheless, this toxicity should be assessed extensively before applying E7107 to UM patients, since these patients either have lost one eye or may have pre-existent visual impairment due to radiation therapy. To optimize its efficacy, E7107 could be used with an altered administration method e.g. nanoparticles for direct delivery to the liver for a higher efficacy [48]. Recently, a phase I clinical trial in myeloid neoplasia, including MDS, was successfully concluded on splicing inhibitor H3B-8800 (chemically similar to E7107) [49]. Similar to E7107, H3B-8800 has also shown a preferential sensitivity towards spliceosome mutant cells [50]. The study demonstrated that this drug has acceptable adverse events and no visual impairment was observed during the study [49]. Interestingly, they have identified a subset of patients who could benefit from this treatment. These patients all had a *SF3B1* mutation. This shows the therapeutic potential of these compounds.

## 5. Conclusions

Further research into the *in vivo* efficacy and toxicity of E7107 and other splicing inhibitors is warranted. Spliceosome inhibitors might be a therapeutic option in spliceosome mutant malignancies including UM. We have demonstrated decreases of cell viability and aberrant transcripts formation of target genes in *SF3B1*^MUT^ UM. This shows that splicing inhibitors such as E7107 can have therapeutic potential in *SF3B1*^MUT^ UM.

## Data Availability

All data produced in the present study are available upon reasonable request to the authors

## Supplementary Materials

The following are available online at www.mdpi.com/xxx/s1, Figure S1. PCA plots and heatmaps of UM RNA-seq samples; Figure S2. RT-PCR analysis of the effects of E7107 treatment on splicing inhibition on UM cell lines; Figure S3. Quantification of preferential sensitivity of UM cells to E7107; Figure S4. Aberrant splicing events per sample; Figure S5. Estimates of the effect of increasing E7107 concentrations on zygosity and DNA copy number of *SF3B1*^WT^ tumor slices; Table S1. Primer sequences; Table S2. RNA sequencing alignment metrics; Table S3. Overview of gene expression data; Table S4. Immunohistochemistry (IHC) settings; Table S5. UM tumor mutational status; Table S6. Alternative splicing normal distribution analysis; Table S7. Decreased aberrant transcripts after E7107 exposure.

## Author Contributions

Conceptualization, A.K. and E.K.; methodology, A.K.; H.J.G.W.; J.R.; E.B. and E.K.; software, T.B. and H.J.G.W.; investigation, J.Q.N.N.; W.D.; A.M.C.H.J.L.; T.B.; T.P.P.B.; resources, R.M.V. and D.P.; data curation, J.Q.N.N.; W.D. and A.M.C.H.J.L.; writing—original draft preparation, J.Q.N.N.; writing—review and editing, W.D.; A.M.C.H.J.L.; T.B.; T.P.P.B.; R.M.V.; H.J.G.W.; J.R.; D.P.; A.K.; E.B. and E.K.; supervision, A.K.; E.B. and E.K.; project administration, A.K.; E.B. and E.K.; funding acquisition, A.K.; E.B. and E.K.

## Funding

This research was funded by Combined Ophthalmic Research Rotterdam (CORR 5.2.0) and Uitzicht (UZ-2018-4).

## Institutional Review Board Statement

The study was conducted according to the guidelines of the Declaration of Helsinki and approved by the Medical ethics committee of the Erasmus Medical Centre (OZR nr 2009-17, MEC-2009-375, 12 November 2009).

## Informed Consent Statement

Informed consent was obtained from all subjects involved in the study.

## Data Availability Statement

All transcriptome count data is in the supporting information files. Our ethics committee does not allow sharing of individual patient or control genotype information in the public domain.

Acknowledgments

We would like to thank Luuk Perdaems, Eva Medico-Salsench and Niels van der Horst for their contribution in the PCR design.

## Conflicts of Interest

The authors declare no conflict of interest. The funders had no role in the design of the study; in the collection, analyses, or interpretation of data; in the writing of the manuscript, or in the decision to publish the results.

## Abbreviations

ARMC9: Armadillo Repeat Containing 9
BAP1: BRCA1 associated protein 1
CDK2: Cyclin-dependent kinase 2
CHMP2A: Charged Multivesicular Body Protein 2A
CNV: Copy Number Variation
CPM: Counts per mean
CYSLR2: Cysteinyl Leukotriene Receptor 2
DLST: Dihydrolipoamide S-Succinyltransferase
DPH5: Diphthamide Biosynthesis 5
EIF1AX: Eukaryotic Translation Initiation Factor 1A X-Linked
EMC7 ER: Membrane Protein Complex Subunit 7
ENOSF1: Enolase Superfamily Member 1
FCS: Fetal Calf Serum
FRASER: Find RAre Splicing Events in RNA_seq
GNA11: G protein Subunit Alpha 11
GNAQ: G protein Subunit Alpha Q
H&E: Hematoxylin and Eosin
IHC: Immunohistochemistry
LC: Lethal concentration
MDS: Myelodysplastic syndrome
MelanA: Melanoma antigen
MIB-1: Mind bomb E3 Ubiquitin Protein Ligase 1
MISO: Mixture-of-Isoforms
NMD: Nonsense-Mediated Decay
PLCB4: Phospholipase C Beta 4
P/S: Penicillin/Streptomycin
RT-PCR: Reverse transcription polymerase chain reaction
*SF3B1*: Splicing factor 3b Subunit 1
SRSF2: Serine- and Arginine-rich splicing factor 2
TBC1D3H: TBC1 Domain Family Member 3H
TMM: Trimmed mean per million
TPM: Transcripts per million
UM: Uveal melanoma

## Notes

### Competing Interest Statement

The authors have declared no competing interest.

### Funding Statement

This study was funded by Combined Ophthalmic Research Rotterdam (CORR 5.2.0) and Uitzicht (UZ-2018-4).

### Author Declarations

Ethics committee/IRB of Erasmus MC gave ethical approval for this work

