## Supplementary material for "Spliceosome Inhibition in *SF3B1*-Mutated Uveal Melanoma": UM E7107 Supplementary CANCERS_rebuttal.docx

Original research

**Citation:** Lastname, F.; Lastname, F.; Lastname, F. Title. *Cancers* **2021**, *13*, x. https://doi.org/10.3390/xxxxx

Academic Editor: Firstname Lastname

Received: date

Accepted: date

Published: date

**Publisher’s Note:** MDPI stays neutral with regard to jurisdictional claims in published maps and institutional affiliations.

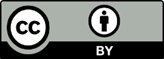

**Copyright:** © 2021 by the authors. Submitted for possible open access publication under the terms and conditions of the Creative Commons Attribution (CC BY) license (https://creativecommons.org/licenses/by/4.0/).

^1^	Department of Ophthalmology, Erasmus MC Cancer Institute, Erasmus MC Medical Center Rotterdam, 3000 CA Rotterdam, The Netherlands.

^2^	Department of Clinical Genetics, Erasmus MC Cancer Institute, Erasmus MC Medical Center Rotterdam, 3000 CA Rotterdam, The Netherlands.

^3^	The Rotterdam Eye Hospital, 3011 BH Rotterdam, The Netherlands.

^4^	Department of Pathology, Section Ophthalmic Pathology, Erasmus MC Cancer Institute, Erasmus MC Medical Center Rotterdam, 3000 CA Rotterdam, The Netherlands.

^5^	Department of Pathology, Leiden University Medical Center, 2333 ZA Leiden, The Netherlands.

^6^	Cancer Computational Biology Center, Erasmus MC Cancer Institute, University Medical Center Rotterdam, 3000 CA Rotterdam, The Netherlands.

^7^	Department of Urology, Erasmus MC Cancer Institute, University Medical Center Rotterdam, 3000 CA Rotterdam, The Netherlands.

^8^	Department of Immunology, Erasmus MC Cancer Institute, University Medical Center Rotterdam, 3000 CA Rotterdam, The Netherlands.

The Rotterdam Ocular Melanoma Study Group (ROMS) is a collaborative research group with members from the Rotterdam Eye Hospital, Departments of Ophthalmology, Pathology and Clinical Genetics, of the Erasmus MC, Rotterdam, the Netherlands

Table of contents

1. Figure S1. PCA plots and heatmaps of UM RNA-seq samples.
2. Figure S2. RT-PCR analysis of the effect of E7107 treatment on splicing inhibition on UM cell lines.
3. Figure S3. Quantification of preferential sensitivity of UM cells to E7107.
4. Figure S4. Aberrant splicing events per sample.
5. Figure S5. Estimates of the effect of increasing E7107 concentrations on zygosity and DNA copy number of *SF3B1*^WT^ tumor slices.
6. Table S1. RT-PCR primers.
7. Table S2. RNA sequencing alignment metrics.
8. Table S3. Overview of gene expression data (excel file)
9. Table S4. Immunohistochemistry (IHC) settings.
10. Table S5. UM tumor mutational status.
11. Table S6. Alternative splicing normal distribution analysis.
12. Table S7. Decreased aberrant transcripts after E7107 exposure (excel file)

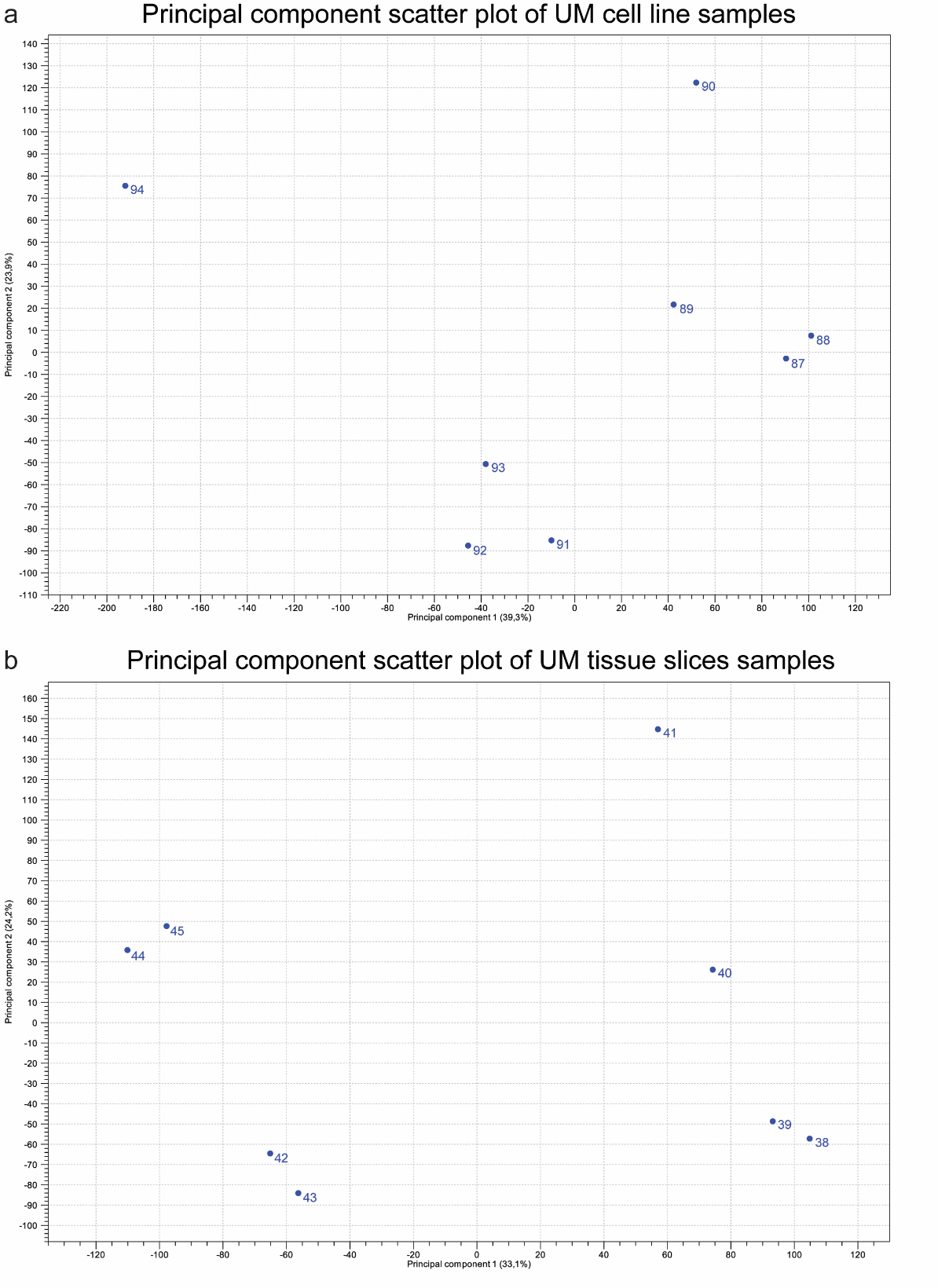

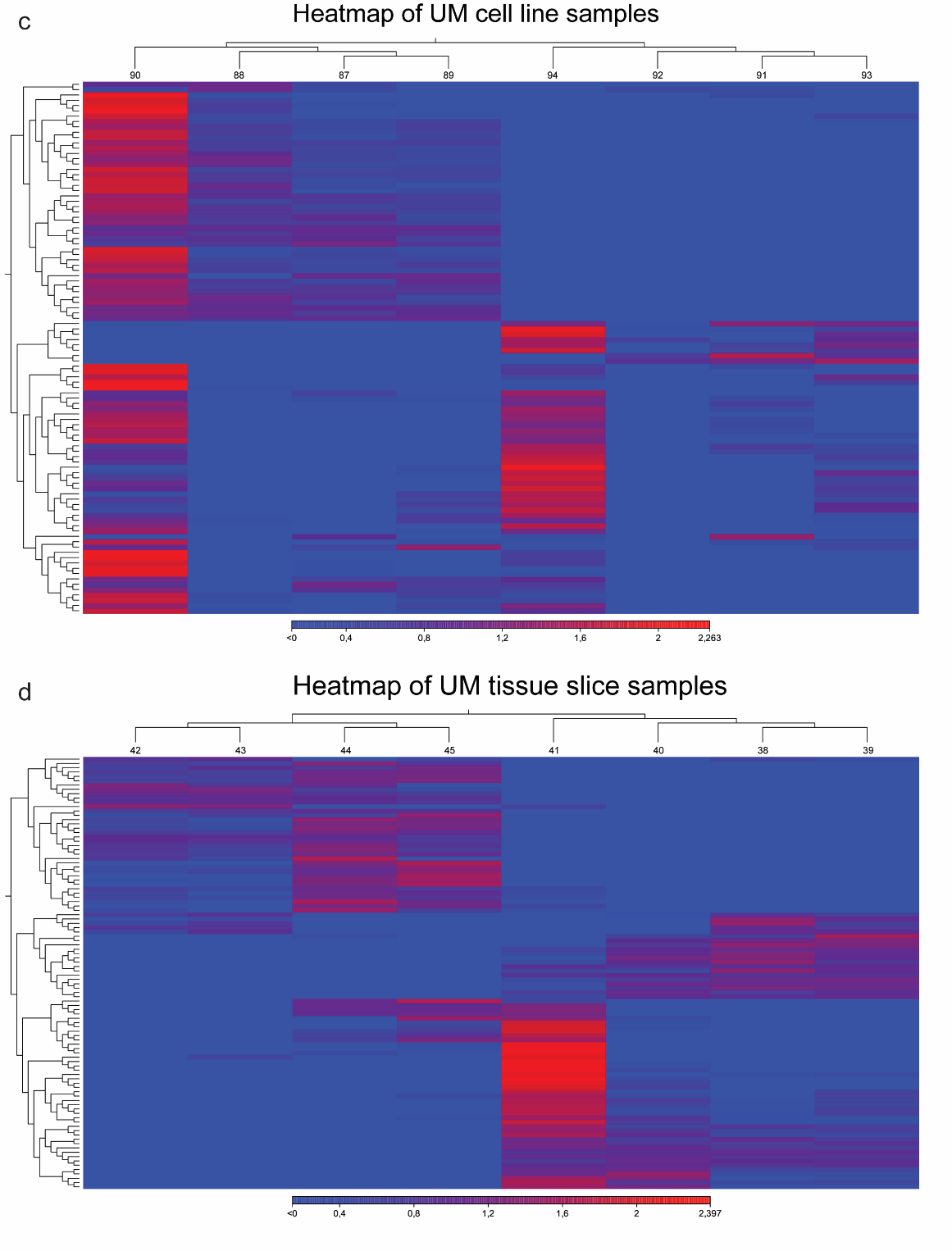
**Figure S1. PCA plots and heatmaps of UM RNA-seq samples.** PCA plots of (a) *SF3B1*^MUT^ and *SF3B1*^WT^ cell line samples and (b) *SF3B1*^MUT^ and *SF3B1*^WT^ tissue slices samples. Heatmaps of (c) *SF3B1*^MUT^ and *SF3B1*^WT^ cell line samples and (d) *SF3B1*^MUT^ and *SF3B1*^WT^ tissue slices samples. Samples 38-41 = *SF3B1*^MUT^ tumor slices; samples 42-45 = *SF3B1*^WT^ tumor slices; samples 87-90 = *SF3B1*^MUT^ cell line; samples 91-94 *SF3B1*^WT^ cell line (conditions see Table S2).

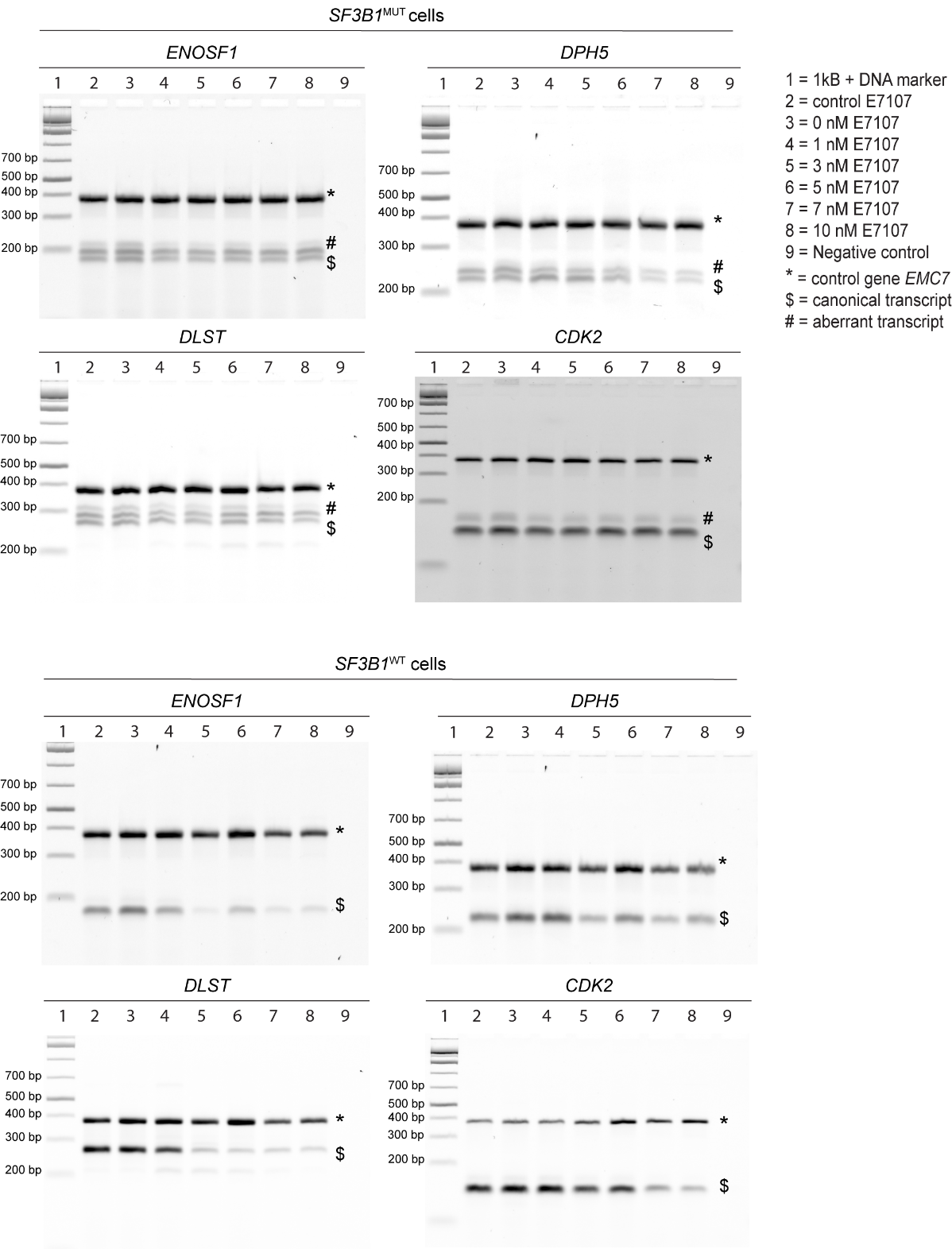
**Figure S2. RT-PCR analysis of the effect of E7107 treatment on splicing inhibition on UM cell lines.** Multiplex RT-PCR products for transcription factors *ENOSF1*, *DPH5*, *DLST* and *CDK2* on *SF3B1*^MUT^ and *SF3B1*^WT^ cells treated with different concentrations of E7107. Amplification of the *SF3B1*^MUT^ sensitive genes resulted in both aberrant and canonical transcript formation for *SF3B1*^MUT^ cells, whereas the *SF3B1*^WT^ products only displayed canonical transcript formation. The insensitive gene *EMC7* was used as a loading control for both cDNA quality and quantity.

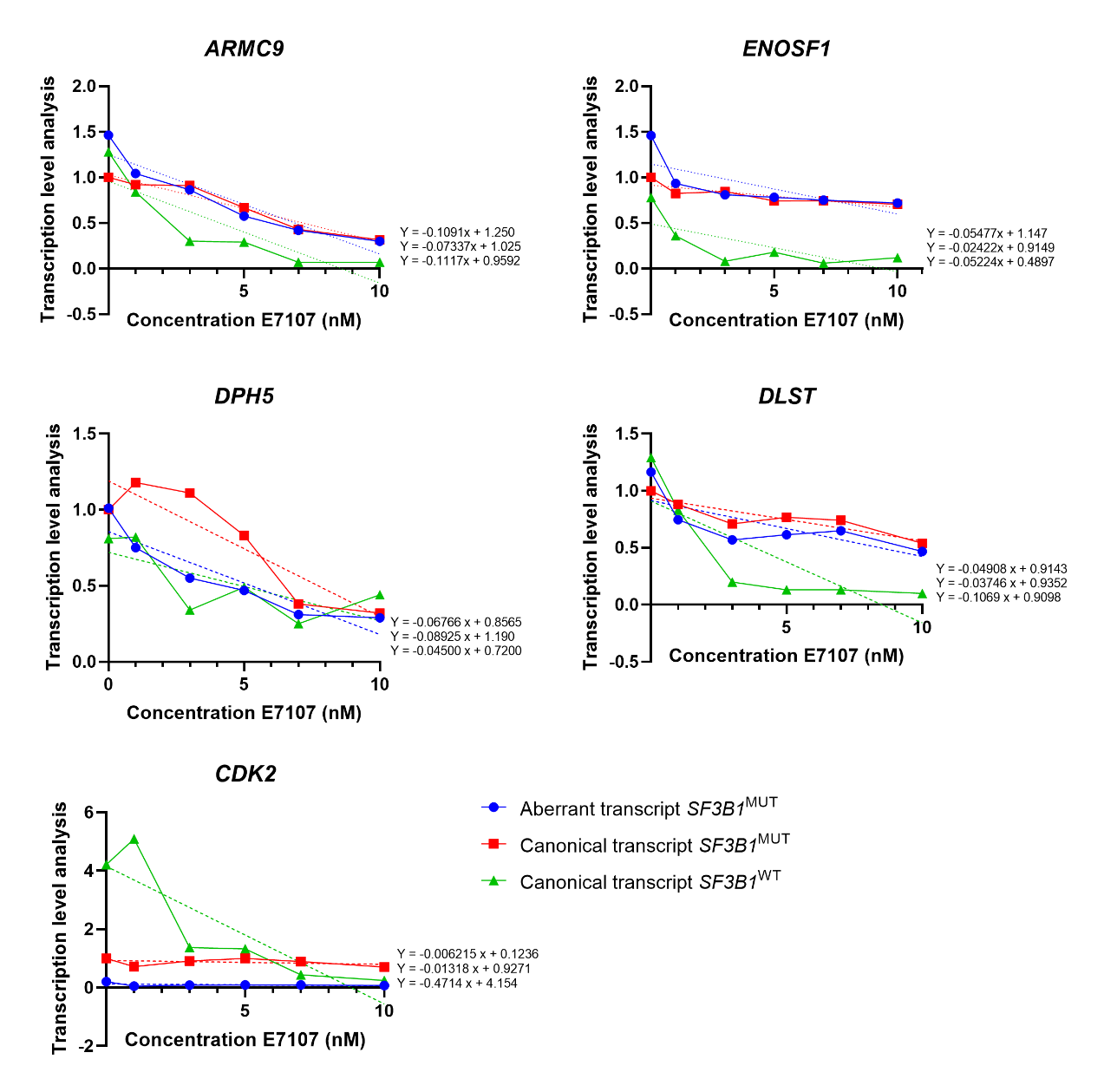

**Figure S3. Quantification of preferential sensitivity of *SF3B1*^MUT^ and *SF3B1*^WT^ UM cells to E7107.** Aberrant and canonical transcripts formation was quantified using ImageQuantTL. Transcription level analysis in UM cells on *ARMC9*, *ENOSF1*, *DPH5*, *DLST* and *CDK2*. Linear regression analysis was used to determine the degree of splicing inhibition (regression comparison between aberrant and canonical transcripts in *SF3B1*^MUT^ UM cells: *ARMC9*, *p* = 0.1195; *ENOSF1*, *p* = 0.2730; *DPH5*, *p* = 0.4093; *DLST*, *p* = 0.6249; *CDK2*, *p* = 0.6989).

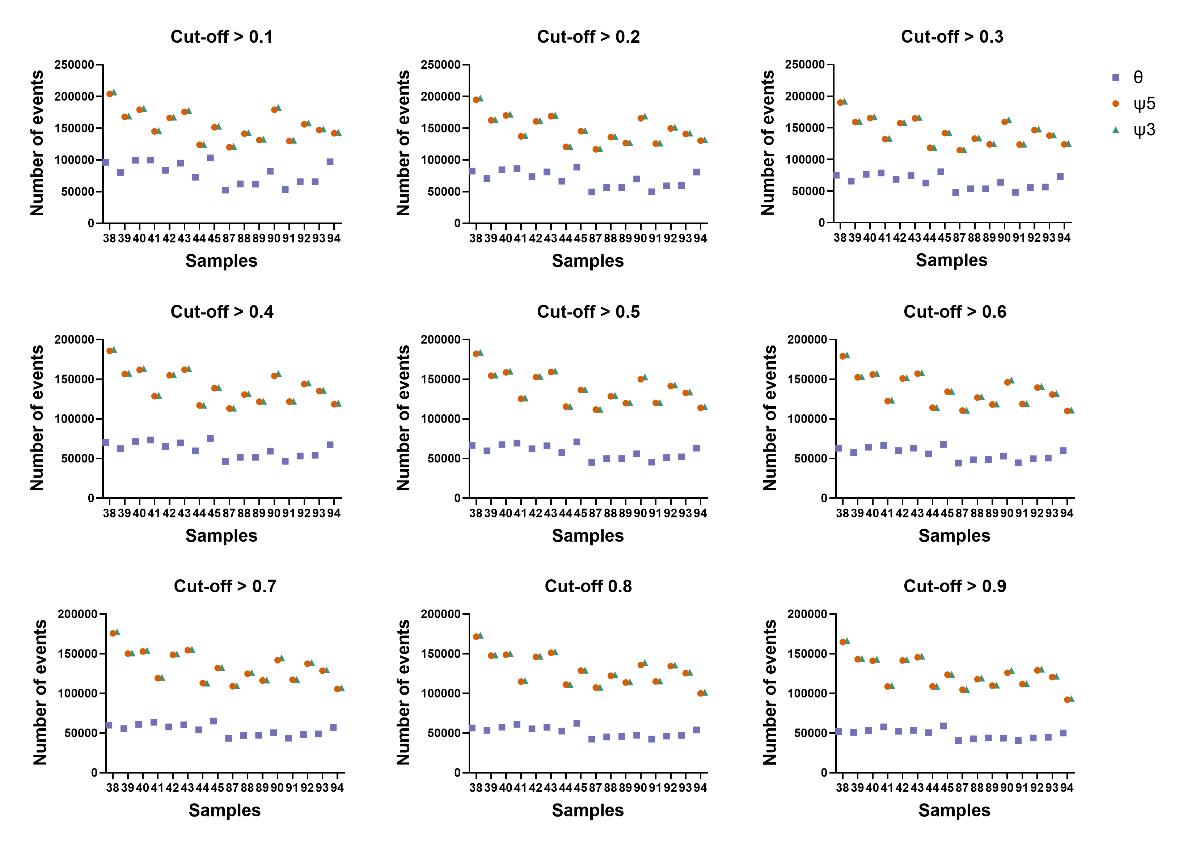

**Figure S4. Aberrant splicing events per sample.** The number of aberrant splicing events per sample, grouped per 0.1 psi (ψ) cut-off values. ψ3 = aberrant donor site (green); ψ5 = aberrant acceptor site usage (orange); θ = intron retention (purple). Samples 38-41 = *SF3B1*^MUT^ tumor slices; samples 42-45 = *SF3B1*^WT^ tumor slices; samples 87-90 = *SF3B1*^MUT^ cell line; samples 91-94 *SF3B1*^WT^ cell line (conditions see Table S2). Per sample (x-axis), the number of events (ψ3/ψ5/θ) observed is depicted (y-axis). Overall, there were more aberrant donor/acceptor site usage events compared to aberrant intron retention per sample.

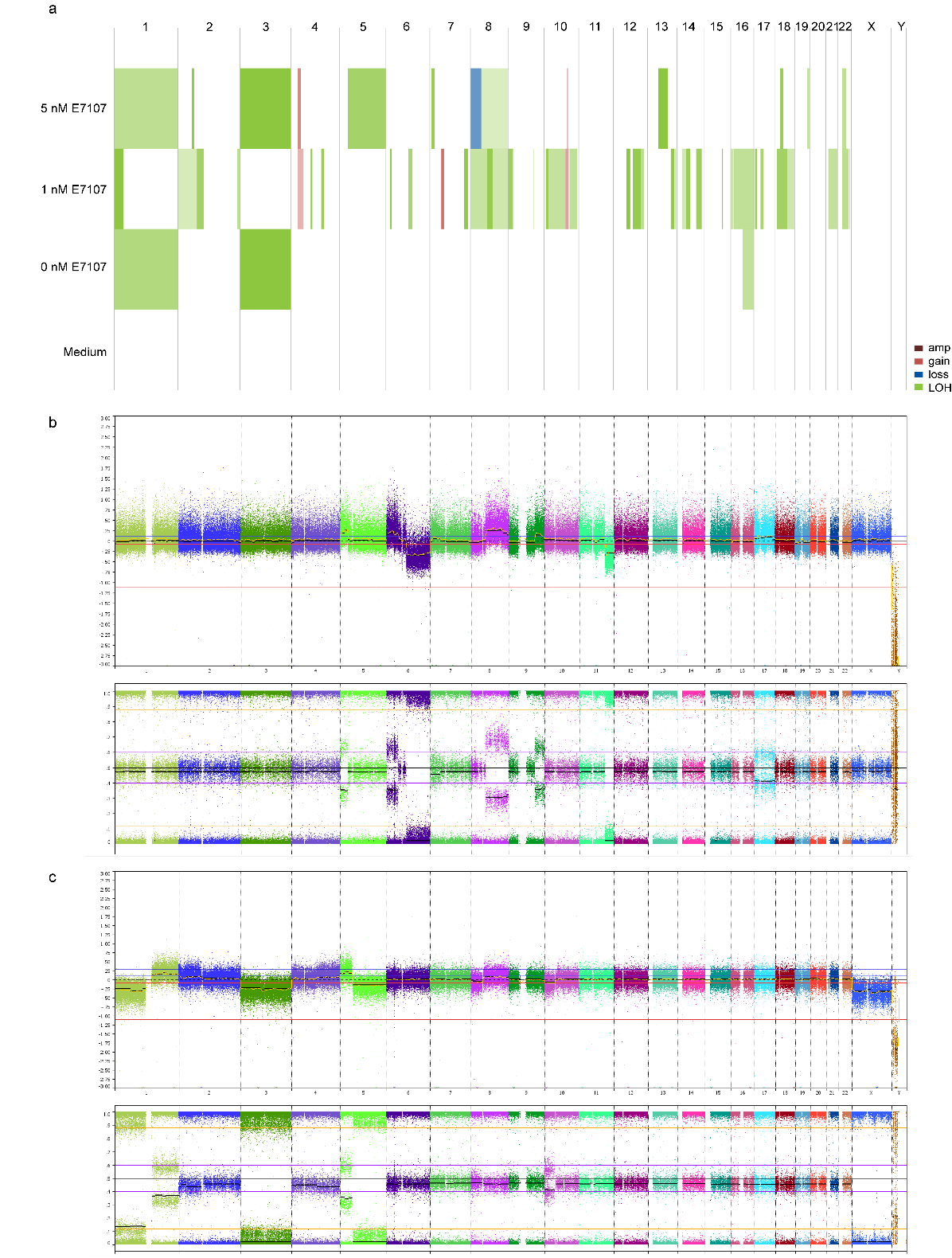

**Figure S5. Estimates of the effect of increasing E7107 concentrations on zygosity and DNA copy number of *SF3B1*^WT^ tumor slices.** (a) The Superfreq DNA copy number and allelic distribution estimates hint at an increasing effect of E7107. An increase in copy number neutral loss of heterozygosity (LOH) is seen after E7107 exposure, especially at 1 nM E7107. Amp, 4+ copies; gain, 3 copies; loss, <2 copies; LOH, 2 copies of the same allele. Each chromosome is depicted on the x-axis. The conditions were normalized to medium. The CNV of the original (b) *SF3B1*^MUT^ and (c) *SF3B1*^WT^ tumor.

**Table S1.** **Primers sequences.**

| **Gene** | **Forward** | **Reverse** |
| --- | --- | --- |
| *ARMC9* | GACATCATGGAAGCCGATCTG | CCTTGTCTTCCCCGTGTTG |
| *ENOSF1* | GATGACATGCGAAGATGCCA | GCTTGAACTTGGCCAGCTT |
| *DPH5* | TGGACAGACACTTGGAGACC | GGGCTGCTTGGTTTACACTC |
| *DLST* | AGTGTCTTCAGTGTTCGCTT | GCTTCAATCACGCCATTTGC |
| *CDK2* | GGATGCCTCTGCTCTCACTG | AATGGCAGAAAGCTAGGCCC |

**Table S2.** **RNA sequencing alignment metrics.** CL = cell line; TS = tumor slice.

| **Sample** | **Condition** | **Source** | **Read counts** | **Paired, mapped pairs (%)** | **Paired, broken pairs (%)** | **Paired, not mapped (%)** | **Mapped to genes (%)** |
| --- | --- | --- | --- | --- | --- | --- | --- |
| 38 | *SF3B1*^MUT^ Medium TS | Tumor 014 | 149,922,012 | 93.99 | 3.39 | 2.63 | 97.57 |
| 39 | *SF3B1*^MUT^ 0 nM E7107 TS | Tumor 014 | 81,519,640 | 94.68 | 3.07 | 2.25 | 97.88 |
| 40 | *SF3B1*^MUT^ 1 nM E7107 TS | Tumor 014 | 101,396,164 | 94.31 | 3.44 | 2.25 | 98.04 |
| 41 | *SF3B1*^MUT^ 5 nM E7107 TS | Tumor 014 | 87,217,358 | 93.24 | 3.78 | 2.99 | 97.35 |
| 42 | *SF3B1*^WT^ Medium TS | Tumor 006 | 107,258,286 | 93.58 | 4.34 | 2.08 | 98.25 |
| 43 | *SF3B1*^WT^ 0 nM E7107 TS | Tumor 006 | 90,112,262 | 95.04 | 2.95 | 2.01 | 98.03 |
| 44 | *SF3B1*^WT^ 1 nM E7107 TS | Tumor 006 | 101,469,750 | 94.03 | 3.75 | 2.22 | 98.29 |
| 45 | *SF3B1*^WT^ 5 nM E7107 TS | Tumor 006 | 90,129,936 | 93.73 | 3.61 | 2.66 | 97.54 |
| 87 | *SF3B1*^MUT^ Medium CL | Mel202 | 71,577,654 | 95.98 | 1.60 | 2.42 | 98.55 |
| 88 | *SF3B1*^MUT^ 0 nM E7107 CL | Mel202 | 87,036,100 | 95.46 | 2.38 | 2.16 | 98.75 |
| 89 | *SF3B1*^MUT^ 1 nM E7107 CL | Mel202 | 72,292,188 | 95.63 | 1.84 | 2.53 | 98.52 |
| 90 | *SF3B1*^MUT^ 5 nM E7107 CL | Mel202 | 169,070,054 | 94.74 | 3.04 | 2.22 | 98.80 |
| 91 | *SF3B1*^WT^ Medium CL | 92.1 | 67,958,438 | 95.60 | 2.02 | 2.37 | 98.74 |
| 92 | *SF3B1*^WT^ 0 nM E7107 CL | 92.1 | 88,471,060 | 95.65 | 2.18 | 2.17 | 98.91 |
| 93 | *SF3B1*^WT^ 1 nM E7107 CL | 92.1 | 99,274,780 | 95.64 | 2.29 | 2.07 | 98.96 |
| 94 | *SF3B1*^WT^ 5 nM E7107 CL | 92.1 | 79,667,126 | 94.11 | 3.12 | 2.78 | 98.12 |

**Table S4.** **Immunohistochemistry (IHC) settings.** AB = antibody.

| **Antibody** | **Type** | **Company** | **Clone** | **Procedure** | **AB incubation time** |
| --- | --- | --- | --- | --- | --- |
| Melan-A | Anti-mouse | Ventana | A103 | Ultraview CC1 36’ | 28 minutes |
| BAP1 | Anti-mouse | Santa Cruz | C-4 | Ultraview CC1 64’ | 32 minutes |
| MIB-1 | Anti-rabbit | Ventana | 30-9 | Ultraview CC1 32’ | 32 minutes |
| Caspase-3 | Anti-rabbit | Ventana | 5A1E | Ultraview CC1 64’ | 32 minutes |

**Table S5.** **UM tumor mutational status.**

| **Tumor** | **Primary driver mutation** | **Secondary driver mutation** | **Tumor location** |
| --- | --- | --- | --- |
| 01 | *GNAQ* (c.626A>T) | *SF3B1* (c.1874G>A; R625L) | choroid |
| 02 | *GNA11* (c.626A>T) | BAP1 IHC neg | choroid |
| 03 | *GNA11* (c.626A>T) | *EIF1AX* (c.25G>C) | choroid |
| 04 | *GNAQ* (c.626A>C) | *EIF1AX* (c.28A>G) | choroid |
| 05 | *GNAQ* (c.626A>T) | # | choroid |
| 06 | *GNA11* (c.626A>T) | *BAP1* (c.1985_1988del) and BAP1 IHC neg | choroid |
| 07 | *GNA11* (c.626A>T) | *BAP1* (c.1754_1755insT) and BAP1 IHC neg | choroid |
| 08 | *GNAQ* (c.626A>C) | *EIF1AX* (c.5C>T) | choroid |
| 09 | *GNAQ* (c.626A>C) | BAP1 IHC neg | ciliary body |
| 10 | *GNA11* (c.626A>T) | *BAP1* (c.68-2A>G) and BAP1 IHC neg | choroid |
| 11 | *PLCB4* (c.1888_1889delinsTT) | *SF3B1* (c.1873C>T; R625C) | choroid |
| 12 | * | *BAP1* (c.1878del) and BAP1 IHC neg | choroid |
| 13 | *GNAQ* (c.626A>T) | *BAP1* (c.1379C>G) and BAP1 IHC neg | choroid |
| 14 | *GNAQ* (c.626A>C) | *SF3B1* (c.1873C>T; R625C) | choroid |
| 15 | *GNA11* (c.626A>T) | *EIF1AX* (c.17-2A>C) | choroid |
| 16 | *GNA11* (c.626A>T) | *EIF1AX* (c.11A>G) | choroid |
| 17 | *GNAQ* (c.626A>T) | *BAP1* (c.1951-1955delinsG) and BAP1 IHC neg | choroid |
| 18 | *GNA11* (c.626A>T) | # | choroid |
| 19 | *GNA11* (c.626A>T) | *BAP1* (c.550G>T) and BAP1 IHC neg | ciliary body |
| 20 | *CYSLTR2* (c.386T>A) | *BAP1* (c.691_705del) and BAP1 IHC neg | ciliary body |

* = wild-type for *GNAQ* and *GNA11*; # = wild-type for *SF3B1* , *EIF1AX* or BAP1 and BAP1 IHC positive.

**Table S6.** **Alternative splicing normal distribution analysis.**

| **Bayes Factor range** | **Counts per cell line sample** | | | | | |
| --- | --- | --- | --- | --- | --- | --- |
|  | 0 nM E7107 *SF3B1*^MUT^ | 1 nM E7107 *SF3B1*^MUT^ | 5 nM E7107 *SF3B1*^MUT^ | 0 nM E7107 *SF3B1*^WT^ | 1 nM E7107 *SF3B1*^WT^ | 5 nM E7107 *SF3B1*^WT^ |
| (0, 10) | 203 | 200 | 94 | 133 | 127 | 111 |
| (10, 100) | 445 | 515 | 257 | 383 | 314 | 270 |
| (100, 1e+3) | 223 | 265 | 150 | 152 | 147 | 126 |
| (1e+3, 1e+4) | 137 | 147 | 98 | 116 | 89 | 76 |
| (1e+4, 1e+5) | 122 | 111 | 63 | 82 | 80 | 61 |
| (1e+5, 1e+6) | 86 | 83 | 62 | 51 | 53 | 52 |
| (1e+6, 1e+7) | 82 | 60 | 48 | 44 | 45 | 35 |
| (1e+7, 1e+8) | 76 | 61 | 52 | 42 | 40 | 31 |
| (1e+8, 1e+9) | 58 | 70 | 33 | 41 | 28 | 36 |
| (1e+9, 1e+10) | 49 | 44 | 35 | 27 | 28 | 25 |
| (1e+10, 1e+11) | 34 | 51 | 27 | 30 | 32 | 30 |
| (1e+11, 1e+12) | 841 | 749 | 1427 | 605 | 723 | 853 |
| Kolmogorov-Smirnov  Test | *p* > 0.05 | | | *p* > 0.05 | | |
| **Bayes Factor range** | **Counts per tumor sample** | | | | | |
|  | 0 nM E7107 *SF3B1*^MUT^ | 1 nM E7107 *SF3B1*^MUT^ | 5 nM E7107 *SF3B1*^MUT^ | 0 nM E7107 *SF3B1*^WT^ | 1 nM E7107 *SF3B1*^WT^ | 5 nM E7107 *SF3B1*^WT^ |
| (0, 10) | 110 | 81 | 44 | 228 | 86 | 146 |
| (10, 100) | 180 | 154 | 94 | 515 | 231 | 328 |
| (100, 1e+3) | 69 | 57 | 42 | 219 | 122 | 165 |
| (1e+3, 1e+4) | 35 | 42 | 34 | 146 | 81 | 118 |
| (1e+4, 1e+5) | 27 | 22 | 18 | 70 | 66 | 83 |
| (1e+5, 1e+6) | 18 | 15 | 23 | 70 | 45 | 75 |
| (1e+6, 1e+7) | 17 | 13 | 6 | 55 | 59 | 52 |
| (1e+7, 1e+8) | 14 | 13 | 7 | 43 | 46 | 44 |
| (1e+8, 1e+9) | 10 | 6 | 14 | 39 | 34 | 48 |
| (1e+9, 1e+10) | 9 | 13 | 8 | 23 | 22 | 28 |
| (1e+10, 1e+11) | 9 | 10 | 7 | 32 | 32 | 30 |
| (1e+11, 1e+12) | 106 | 178 | 307 | 363 | 979 | 686 |
| Kolmogorov-Smirnov  Test | *p* > 0.05 | | | *p* > 0.05 | | |
